# Eating Disorders and Parkinson’s Disease - 2: Population Burden, Genetic Epidemiology and Shared Genomics

**DOI:** 10.64898/2026.01.03.26343328

**Authors:** Andrew W Bergen, Carolina Makowski, Michael Garvin, Gulcan Cil, Angeline Krueger, Karlee McGlone, Irene Litvan, Walter F Kaye

## Abstract

**Objective:** We reviewed the epidemiologic and genomic literature and performed genomic analyses to identify population burdens (Eating Disorders, ED and Parkinson’s Disease, PD) and shared genetic risk (Anorexia Nervosa, AN and PD), after we previously demonstrated two-to four-fold relative risks of a family history of Parkinson’s Disease in families of individuals with ED.

**Method:** We reviewed the epidemiology of both disorders and published genome-wide association studies (GWAS), searched PD GWAS findings with AN associated genome-wide significant SNPs and associated genes and regions, and performed conditional/conjunctional false discovery rate genomic analyses of AN and PD summary statistics to identify shared genetics.

**Results:** The global population burden of ED and PD are similar despite differences in age of onset and sex ratio. GWAS review and linkage disequilibrium analysis showed that genome-wide significant variants from AN GWAS and PD GWAS in the chr3p21.31 region are correlated. We identified a chr3p21.31 variant with joint association with both disorders; this variant has functional linkages to 40 genes.

**Conclusions:** ED and PD share neuropsychological, neurobiological, and genetic risk factors, including association with the complex chr3p21.31 locus. Translational analyses leveraging disorder-specific research resources may benefit our understanding of the genetics and neuropsychiatric mechanisms of both disorders.

**Highlights:**

- ED and PD have similar age-standardized disability life years (AS-DALYS); the burden of both disorders are expected to increase in the future.
- Genomic epidemiology literature review reveals AN and PD genetic architectures are highly polygenic (AN >> PD), and, AN variant discoverability is less than PD variant discoverability.
- AN and PD are jointly associated at rs1352420 at chr3p21.31, a region with multiple AN and PD comorbid and genetically correlated trait associations.
- Cross-disorder research offers opportunities to identify shared variants and explore mechanistic hypotheses

## Introduction and Aims

We recently demonstrated in a clinical study that families with an eating disorder (ED) have two to four-fold relative risks of a family history of Parkinson’s disease (FHoPD) compared to families without an ED (Krueger, Angeline et al. submitted 2025). Despite differences in age at onset and peak prevalence, sex ratios and diagnostic symptoms, Anorexia Nervosa (AN) and Parkinson’s Disease (PD) share traits in multiple domains: temperament, e.g., harm avoidance; psychological, e.g., anxiety; metabolic, e.g., weight loss; and neurobiologic, e.g., dopaminergic alterations. See Krueger (Krueger, Angeline et al. submitted 2025) for details.

Stimulating cross-disorder research to unravel shared genetic and neurobiological mechanisms will benefit translational research for both disorders. To investigate and compare AN and PD epidemiology, genetic epidemiology, genetics and potential common mechanisms, we: reviewed published epidemiologic and genetic epidemiologic literature; examined genome-wide association study (GWAS), cross-disorder and mechanistic literature; and, performed novel genomic analyses of AN and PD summary statistics.

## Methods

We reviewed the Global Burden of Diseases, Injuries and Risk Factors Study (GBD) mental health and neurological disorders literature to compare ED and PD burden estimates to provide translational context for shared genetics. We searched genetic and genomic epidemiology literature of AN, PD and psychiatric-neurologic disorders for selected measures. We selected the published AN GWAS (Watson et al. 2019; Duncan et al. 2017), extracted genome-wide significant (GWS) SNPs, assigned and gene-wise significant genes and author-defined associated regions, and searched the NHGRI-EBI GWAS catalog (Cerezo et al. 2025) for overlapping PD GWS associations. We searched the literature for mechanistic associations using AN assigned and gene-wise significant genes and PD’s Medical Subject Heading. We used LDlink’s LDpair tool and 1000 Genomes HapMap data from 503 European ancestry individuals to estimate linkage disequilibrium (LD) and the UCSC Genome Browser for annotation (Perez et al. 2025; Machiela and Chanock 2015).

We carried out conditional/conjunctional false discovery rate (cFDR) analyses (Smeland et al. 2020) to identify overlapping genetic variants between AN and PD (Watson et al. 2019; Chang et al. 2017) using the pleioFDR tool (https://github.com/precimed/pleiofdr). Specifically, we conditioned the GWAS summary statistics for one trait onto another in both directions, where the highest of the resulting pair of FDR values per genetic variant was used to indicate strength of evidence for shared effects between the two traits. We used default settings and an FDR threshold of 0.05 for whole-genome significance, as recommended. Resulting genetic variants and those in LD (r^2^>0.6) were mapped to genes in FUMA (Watanabe et al. 2017) using positional, gene expression (eQTL), and chromatin interaction information.

## Results

### Population Burden

GBD 2019 AS-DALY rate estimates (per 100,000 person years) were 86 (49-137) for EDs (38 (24-56) for AN and Bulimia Nervosa (BN), 10 (4-20) for Binge-Eating Disorder, BED, and 39 (20-67) for Other specified feeding or eating disorder, OSFED) and 80 (73-87) for PD (Ding et al. 2022; Santomauro et al. 2021). ED AS-DALY burden estimates are higher among females (2.4-fold) for ED (peak prevalence in the third decade for women, the fourth decade for men) and higher among males (1.8-fold) for PD (peak prevalence in the eighth and ninth decades) (Ding et al. 2022; Santomauro et al. 2021). Population burdens are highest in high sociodemographic index countries with US, Canada, Asia Pacific, and Western Europe AS-DALY rates twice the global average for ED (Liu et al. 2025) and US, Canada, Argentina, and Central Europe AS-DALY rates twice the global average for PD (Dorsey et al. 2018).

### Genetic Epidemiology

See **Table 1** for precise genetic epidemiologic parameter estimates and uncertainty intervals. AN and PD have similar ∼four-fold familial 1° relative risks (Steinhausen et al. 2015; Thacker and Ascherio 2008). Twin analyses suggest additive genetics account for the majority of AN and BN risk but only ∼one-third of male PD and ∼one-fifth of female PD risk with environment accounting for twice the level of risk (∼two-thirds) in PD, versus AN (∼one-third) (Bulik et al. 2006; Wirdefeldt et al. 2011). Additive common variant genetic heritability estimates of AN are significant and similar to the average psychiatric disorder heritability, whichever method is used (Smeland et al. 2025). Common variant heritability estimates of PD are significant but less than the average heritability of other neurologic disorders (which are lower than psychiatric disorders) (Smeland et al. 2025).

**TABLE 1.** Genetic epidemiology of Anorexia Nervosa & Parkinson’s Disease.

| Measure | Measure detail | AN Estimate(s) (95CI) | Sources | Measure detail | PD Estimate(s) (95%CI) | Sources |
| --- | --- | --- | --- | --- | --- | --- |
| Familiality<br>CFRR=Crude<br>Familial Relative Risk<br>A1°R=all 1° relatives<br>F1°R=1° female<br>relatives | CFRR, AN, A1°R of AN<br>proband, national (DK)<br>and<br>CFRR, AN, F1°R of AN<br>proband<br>and<br>CFRR, ED, in families,<br>clinic (UCLA) | CFRR AN, A1°R=3.71<br>(2.35-5.87)<br>and<br>CFRR AN, F1°R=10.97<br>(1.48-229.00)<br>and<br>CFRR ED=4.90 (2.38-10.66) | (Steinhausen et al.<br>2015)<br>Table 2<br>and<br>(Strober et al. 2000),<br>Table 2<br>and<br>Table 3 | Meta-analytic random-<br>effects RR for PD with<br>1° relatives with PD<br>using all studies<br>and<br>by age of onset | Child-par.=2.7 (2.0-3.7)<br>Sib=4.4 (3.1-6.1)*<br>1° rel.=3.8 (3.0-5.0)<br>*p=.049 sib vs child-par.<br>and<br>Early=4.7 (3.2-6.8)<br>Late=2.7 (1.9-3.9)**<br>All=3.7 (2.8-4.8)<br>**p=.041 early vs late | (Thacker and Ascherio<br>2008)<br>Table 3<br>and<br>Table 2 |
| Twin Heritability | Structural equation<br>estimates for additive<br>genetics (a<sup>2</sup>), shared<br>(c<sup>2</sup>) and non-shared (e<sup>2</sup>)<br>environment | a<sup>2</sup>=0.56 (0.00-0.87),<br>c<sup>2</sup>=0.05 (0.00-0.64),<br>e<sup>2</sup>=0.38 (0.13-0.84)<br>both sexes | (Bulik et al. 2006)<br>Swedish Twin Registry<br>Abstract and Table 5 | Structural equation<br>a<sup>2</sup>, c<sup>2</sup>, and e<sup>2</sup> estimates | a<sup>2</sup>=0.34, c<sup>2</sup>=0.00,<br>e<sup>2</sup>=0.66<br>male twins,<br>a<sup>2</sup>=0.19, c<sup>2</sup>=0.12,<br>e<sup>2</sup>=0.69<br>female twins | (Wirdefeldt et al. 2011)<br>Swedish Twin Registry,<br>95%CI not reported<br>Table 4 |
| Common variant<br>(SNP) heritability<br>(h<sup>2</sup>g) on the observed<br>scale and general<br>comment on<br>case:control<br>ascertainment | LDSC h<sup>2</sup>g clinical and<br>population cases and<br>hospital and population<br>controls<br>LDSC h<sup>2</sup>g<br>with clinical, biobank<br>& community cases &<br>controls<br>and<br>MiXeR h<sup>2</sup>g with<br>clinical, biobank &<br>community cases &<br>controls | h<sup>2</sup>g (SE) = 0.172 (0.023)<br>h<sup>2</sup>g (SE) = 0.135 (0.007)<br>and<br>h<sup>2</sup>g (SD) = 0.217<br>(0.004) | (Anttila et al. 2018;<br>Duncan et al. 2017)<br>and<br>(Watson et al. 2019;<br>Smeland et al. 2025)<br>Smeland Table 1<br>and<br>Table S1 | LDSC h<sup>2</sup>g, clinical &<br>familial cases and<br>clinical & population<br>controls<br>LDSC h<sup>2</sup>g<br>and<br>MiXeR h<sup>2</sup>g<br>with clinical, familial,<br>self-reported & proxy (1°<br>relative) cases,<br>and<br>clinical, population &<br>self-reported controls | h<sup>2</sup>g (SE) = 0.105 (0.017)<br>h<sup>2</sup>g (SE) = 0.044 (0.004)<br>and<br>h<sup>2</sup>g (SD) = 0.079<br>(0.001) | (Anttila et al. 2018;<br>International Parkinson<br>Disease Genomics<br>Consortium et al. 2011)<br>and<br>(Nalls et al. 2014;<br>Chang et al. 2017; Nalls<br>et al. 2019; Smeland et<br>al. 2025)<br>Smeland Table 1<br>and<br>Table S1 |
| Polygenicity (π<sub>1</sub>),<br>proportion of causal<br>variants<br>Number causal<br>variants (Nc) explain<br>90% variance | Mean (stdev) of (π<sub>1</sub>)<br>Nc<br>(Holland et al. 2020) | 2.37E-03 (1.51E-04)<br>7983 (523) | (Watson et al. 2019;<br>Smeland et al. 2025)<br>Smeland Table S8 | Mean (stdev) of (π<sub>1</sub>)<br>Nc<br>(Holland et al. 2020) | 1.40E-04 (5.27E-05)<br>867 (47) | (Nalls et al. 2014;<br>Chang et al. 2017; Nalls<br>et al. 2019; Smeland et<br>al. 2025)<br>Smeland Table S1 |
| Discoverability (σ<sup>2</sup><sub>β</sub>),<br>Effect size variance<br>per causal variant | Effect size variance<br>of causal variants<br>(σ<sup>2</sup><sub>β</sub>)<br>(Holland et al. 2020) | 4.20E-05 (2.69E-06) | (Watson et al. 2019;<br>Smeland et al. 2025)<br>Smeland Table S2 | Effect size variance<br>of causal variants<br>(σ<sup>2</sup><sub>β</sub>) (Holland et al.<br>2020) | 1.40E-04 (6.95E-06) | (Nalls et al. 2014;<br>Chang et al. 2017; Nalls<br>et al. 2019; Smeland et<br>al. 2025)<br>Smeland Table S2 |

Using psychiatric and neurologic disease GWAS SNP z-scores and a common reference panel of ∼11 million SNPs, Smeland and colleagues estimated polygenicity, causal variant N (Nc), and discoverability and found mean psychiatric disorder polygenicity and Nc one order of magnitude larger than mean neurologic disorders estimates, while mean discoverability was an order of magnitude smaller in psychiatric than neurologic disorders (Smeland et al. 2025). Estimates based on AN (Watson et al. 2019) and PD (Nalls et al. 2014; Chang et al. 2017; Nalls et al. 2019) GWAS found AN polygenicity and Nc estimates to be ten-fold greater than PD estimates, while AN SNP discoverability was 40% of PD SNP discoverability (Smeland et al. 2025).

Bivariate LDSC identified significant genetic correlation between disorders (Smeland et al. 2025). Polygenicity analysis using bivariate MiXer found a similar correlation and mean (SD) number of AN-associated, PD-associated and shared causal variants [7563 (482); 447 (168), and 420 (148)], with 72% of shared variants estimated to have concordant effects (Smeland et al. 2025).

### Comparison of AN & PD GWAS findings

We identified one AN GWAS locus finding with a GWS PD association at chr3p21.31 (**Table 2**). The chr3p21.31 AN GWS SNP (rs9821797, chr3:48,718,253, *NCKIPSD* IVS5) was the lead SNP in a region (SNPs with *P*<1E-5 and *r*^2^>0.1 with rs9821797) spanning 3.79 Mbp with 96 protein-coding genes; *NCKIPSD* was the nearest protein-coding gene (Watson et al. 2019). The region included the PD GWS SNP rs12497850 (chr3:48,748,989, *IP6K2* IVS1), 30 Kbp distal of rs9821797, assigned to *IP6K2* by QTL analysis (Chang et al. 2017; Nalls et al. 2019). These SNPs are in linkage disequilibrium (*D*’=0.92 and *r*^2^=0.23, *χ*^2^=235.1, *p*<.0001); the AN risk allele (A) is correlated with the PD protective allele (G).

**Table 2.** Genome-wide Significant (GWS) Loci for AN, AN & cross-disorder associations, and shared GWS Loci for PD.

| SNPs, Gene(s) and/or LD Region* | Protein Name | AN or AN cross trait association at SNP, Gene(s) or LD region | Search EMBL GWAS catalog for: SNP, Gene or LD region & "Parkinson" |
| --- | --- | --- | --- |
| SNP rs4622308, chr12:56075401;<br>Genes (all gene-wise significant):<br>chr12q13.2-13.3: *PMEL*, *CDK2*, *RAB5B*,<br>*SUOX*, *IKZF4*, *RPS26*, *ERBB3*,<br>*ENSG00000257411*, *PA2G4*, *RPL41*,<br>*ZC3H10*, *ESYT1*, and,<br>*chr12q14.1: TAFA2/FAM19A2* | Premelanosome protein,<br>Cyclin dependent kinase 2,<br>RAB5B, member RAS oncogene family,<br>Sulfite oxidase,<br>IKAROS family zinc finger 4,<br>Ribosomal protein S26,<br>Erb-b2 receptor tyrosine kinase 3,<br>Proliferation-associated 2G4,<br>Ribosomal protein L41,<br>Zinc finger CCCH-type containing 10,<br>Extended synaptotagmin 1,<br>TAFA Chemokine Like Family Member 2<br>(formerly, Family with sequence similarity<br>19 member A2) | rs4622308 with AN (Duncan et al. 2017) | No PD association with SNP reported.<br>No PD associations with 13 genes reported.<br>No LD region reported. |
| SNPs rs9821797, chr3:48718253<br>(*NCKIPSD* IVS5) (*nearest gene, 110 other*<br>*genes identified, 95 protein-coding genes*);<br>2nd locus rs73088112, chr3:49413352<br>*RHOA* IVS1, LD region includes both<br>SNPs: chr3:47588253-51368253** | NCK interacting protein with SH3 domain | rs9821797 with AN (Watson et al. 2019)<br>and AN-EA (Chen et al. 2023);<br>within LD region: rs13093385, rs13096760,<br>rs73073015, rs9832454 with AN-EA (Chen<br>et al. 2023); rs9873726, AN-MDD (Peyrot<br>and Price 2021); | Within LD region chr3:47588253-51368253,<br>rs12497850 (*IP6K2* IVS1 variant)<br>associated with PD at: 9E-9 (Chang et al.<br>2017) and 1E-10 (Nalls et al. 2019), and<br>3E-9 (Kim et al. 2024).<br>*IP6K2* selected by functional analysis<br>(Chang et al. 2017; Nalls et al. 2019). |
| rs6589488, *CADM1* (*nearest, 1 other*)<br>chr11:114997256-115424956, | Cell Adhesion Molecule 1 | (Watson et al. 2019) | No EMBL GWAS catalog association w/PD |
| rs2287348, chr2: 53881813-54362813,<br>*ASB3*, *ERLEC1* (*nearest, 13 other genes*) | ankyrin repeat and SOCS box containing 3,<br>endoplasmic reticulum lectin 1 | (Watson et al. 2019) and MDD vs AN<br>(Peyrot and Price 2021) | No EMBL GWAS catalog association w/PD |
| rs2008387, chr10: 131269764-131463964,<br>*MGMT* (*nearest, 2 other*) | O-6-Methylguanine-DNA Methyltransferase | (Watson et al. 2019) | No EMBL GWAS catalog association w/PD |
| rs9874207, chr3: 70670750-71074150<br>*FOXP1* (*nearest, 2 other*) | Forkhead box protein 1 | (Watson et al. 2019) | No EMBL GWAS catalog association w/PD |
| rs10747478, chr1: 96699455-97284455<br>*PTBP2* (*nearest, 2 other*) | Polypyrimidine Tract Binding Protein 2 | (Watson et al. 2019) | No EMBL GWAS catalog association w/PD |
| rs370838138, chr5: 24945845-25372845<br>*CHD10* (*nearest*) | Cadherin 10 | (Watson et al. 2019) | No EMBL GWAS catalog association w/PD |
| rs13100344, chr3:93968107-95059107,<br>*NSUN3* (*nearest, 2 other*) | NOP2/Sun RNA methyltransferase 3 | (Watson et al. 2019) | No EMBL GWAS catalog association w/PD |

\*(Watson et al. 2019) defined the LD regions as the basepair region for each newly associated GWS locus (Table 1) for SNPs with $P < 1E-5$ and linkage-disequilibrium $r^2 > 0.1$ with most associated "lead" SNP; nearest gene as nearest gene within the region of LD with lead variant ( $r^2 > 0.6$ +/- 500Kb); and, other genes by location, adult brain eQTL, regulatory chromatin interactions and MAGMA gene-wise analysis (but did not list other genes). \*\*LD region includes LD defined by both rs9821797 and rs73088112.

### ConjFDR Results

We identified one genetic variant (rs1352420, chr3:48,757,291, ∼3 kbp 5’ to *IP6K2*) shared by AN and PD with cFDR analysis (**Table 3**, **Figure 1**). The cFDR SNP mapped to 40 genes using FUMA where four genes (*NCKIPSD*, chr3:48,711,277-48,723,348; *P4HTM*, chr3:49,027,674-49,044,586; *WDR6*, chr3:49,044,824-49,053,384; and *DALRD3*, chr3:49,052,921-49,056,013) were mapped by all three methods (positional, eQTL and chromatin interaction mapping) (**Supplementary Table 1**). The cFDR SNP was linked to *IP6K2* by FUMA by two methods, is 8.3kbp distal and in complete LD (*D*’=1.0 and *r*^2^=1.0; *χ*^2^=1,006.0, *p*<.0001) with rs12497850 (the PD SNP), and is 39kbp distal with rs9821797 (the top AN SNP).

**Figure 1.**
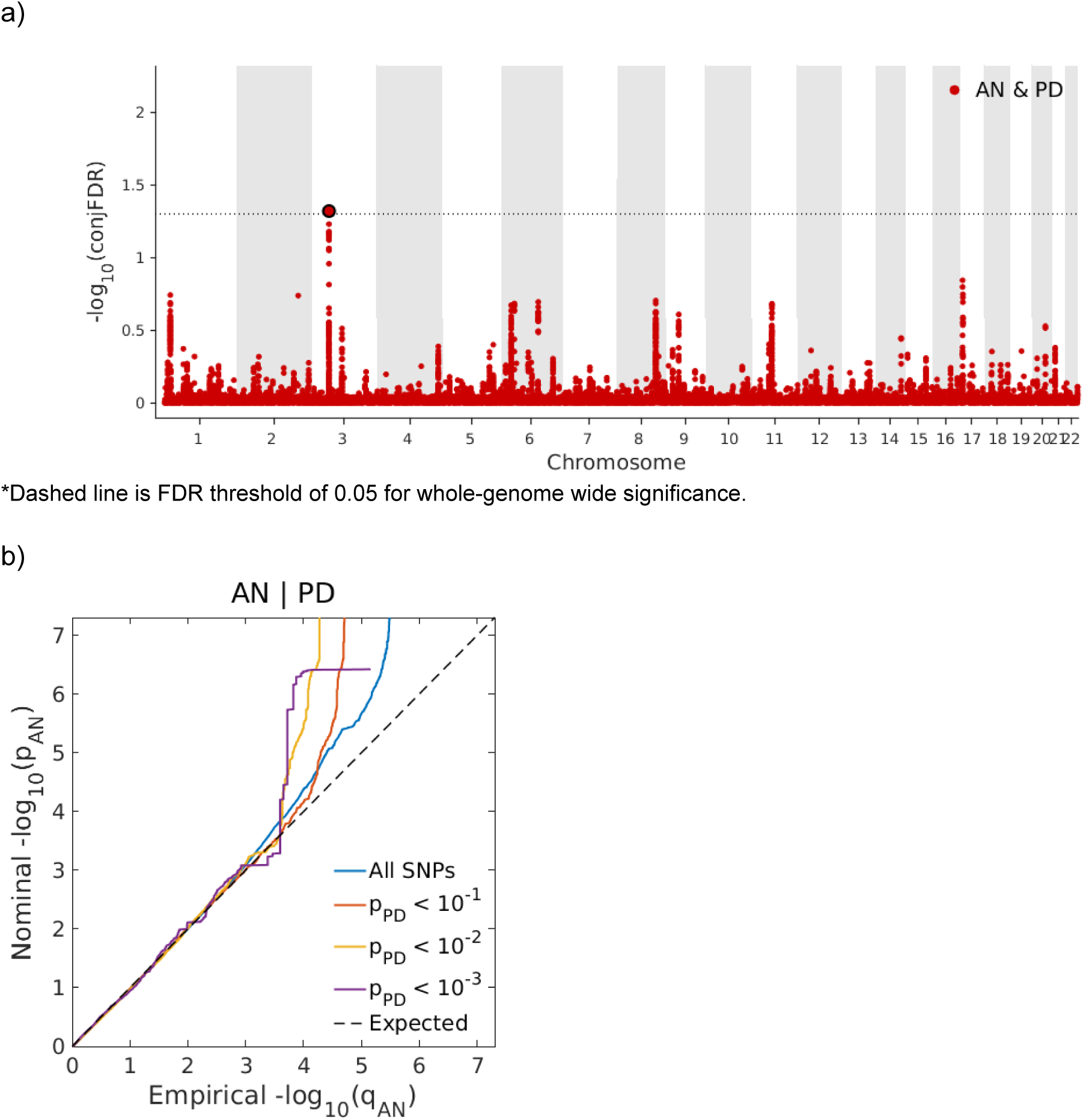
Results of conjFDR analysis of AN and PD: a) Manhattan plot*, and b) QQ plot.

**Table 3.** conjFDR results of overlapping SNP between AN & PD, based on GWAS summary statistics.

| SNP | Chr:coor | beta_AN | se_AN | p_AN | beta_PD | se_PD | p_PD | p_AN condPD | p_PD condAN | fdr_AN | fdr_PD | conjfdr_p_AN_PD |
| --- | --- | --- | --- | --- | --- | --- | --- | --- | --- | --- | --- | --- |
| rs1352420 | 3:48,757,291 | 0.0737 | 0.014 | 1.38E-07 | 0.0760 | 0.0199 | 1.30E-04 | 6.74E-07 | 1.67E-04 | 1.32E-02 | 1.44E-01 | 4.77E-02 |

AN GWAS (Watson et al. 2019); PD GWAS (Watson et al. 2019; Chang et al. 2017). In each analysis (AN and PD), rs1352420 Allele 1=G, Allele 2=A.

## Discussion

ED and PD are disorders with complex genetic and environmental etiologies. Early nosology applied to movement disorder patients, the study of antipsychotic agents and induced Parkinsonism, and secondary Parkinsonism due to the substantia nigra dopaminergic neuron poison MPP+ provided catalysts for etiologic and therapeutic research (Goetz 2011). Positional gene mapping and identification of Lewy body α-synuclein protein aggregates (Polymeropoulos et al. 1997; Spillantini et al. 1997) led to gene (Nalls et al. 2019), brain cell (Bryois et al. 2020) and organelle and molecular process (Michel et al. 2016) discovery in PD. EDs, while at an earlier stage of research in terms of gene discovery, have uncovered genomic (Watson et al. 2019; Hübel et al. 2021), metabolic (Watson et al. 2019; Hebebrand et al. 2025) and neurobiological (Frank et al. 2019) factors associated with the spectrum of behavioral pathologies in EDs and closely related psychiatric disorders, e.g., OCD (Levinson et al. 2019; Yilmaz et al. 2020). Our observations that ED families carry increased relative risk of a PD family history and that AN and PD share psycho-neuro-metabolic features (Krueger, Angeline et al. submitted 2025) and Smeland’s first observations of genetic correlation (Smeland et al. 2023) prompted our cross-disorder research.

Comparing ED and PD epidemiology is challenging due to differences in age of onset, changing epidemiologic environments, and assessment and coding issues for these complex disorders. However, the ongoing global epidemiology project (GBD 2019) estimated similar population burdens, with larger uncertainty for EDs. Recent modeling suggests increasing ED incidence rates, especially in high social development index (SDI) countries, pointing to increasing future ED burden (Liu et al. 2025). Population aging will increase incident and prevalent PD and its population burden; male PD burden is expected to continue to exceed female PD burden (Ding et al. 2022). These trends suggest increasing population burdens of both groups of disorders.

Our review of genetic epidemiologic measures compared AN and PD familial, twin and common variant measures of heritability, polygenicity, Nc and discoverability. Familialities are similar between the disorders overall with greater familiality for female 1° relatives in AN and for early onset in PD. Measures of heritability reinforce the complex nature of both disorders (additive genetics, common environment and unique environment contribute to both disorders), with additive genetic risk greater in AN than in PD, and with unique environment risk greater in PD. Common variant heritability estimates align AN’s genetic risk with other psychiatric diseases, and estimate reduced common variant heritability for PD below that of other neurologic diseases (Smeland et al. 2025). The heterogeneity of PD’s clinical presentation, pathologic findings and increased genetic findings suggests that common, rare and structural genetic variants and environmental risk factors combine to create a disease spectrum (Ye et al. 2023); this complexity may account for increased twin heritability of early onset PD (Goldman et al. 2019) but reduced common variant heritability.

The chr3p21.31 AN:PD conjoint locus is associated with multiple cognitive and neurobiologic phenotypes of interest to both psychiatry and neurology. The top AN SNP was also associated antagonistically (opposite in direction) with educational attainment (EA) in cross-trait analysis (Chen et al. 2023), with additional cross-disorder (with EA and major depression) (Chen et al. 2023; Peyrot and Price 2021) and conjoint (with BMI, age at menarche, schizophrenia, bipolar, major depression, mood instability, neuroticism, and intelligence) (Lu et al. 2024; Breton and Kaufmann 2025; Bang et al. 2023) associations observed in the chr3p21.31 region. The top AN SNP, the LD region and multiple genes in the region have evidence for recent positive selection in the human lineage (Breton and Kaufmann 2025). The top PD SNP in the chr3p21.31 region was associated with neurogenic orthostatic hypotension, cognitive impairment at motor symptom onset, and cognitive decline in PD patients, and with expression of *AMT*, *GMPPB* and *WDR6* in multiple brain regions (Chevalier et al. 2023; Cao et al. 2024). The chr3p21.31 region contains additional GWAS findings for anthropomorphic measures, anxiety, EA, major depression, metabolic measures, neuroticism, physical activity and schizophrenia; all are AN-related traits previously genetically correlated with AN (Duncan et al. 2017; Watson et al. 2019). EA and cognitive function GWAS findings in the LD region are PD-related traits previously genetically correlated with PD (Nalls et al. 2019). The linked genes in the chr3p21.31 region include genes that code for mitochondrial proteins suggesting a role for genes in this region in mitochondrial dysfunction in both AN and PD.

The AN-PD conjointly associated SNP (rs1352420) is the first SNP to be described from hundreds of expected conjointly associated variants (Smeland et al. 2025). The AN GWAS nominated *NCKIPSD* and the PD GWAS nominated *IP6K2*; the AN-PD conjointly associated SNP is linked to both genes by functional criteria and in LD with the AN and the PD top SNPs. *NCKIPSD* was among the top 0.5% of genes associated with functional neuroimaging maps related to “conditioning”, “fear” and “reward” (Murray et al. 2023); *NCKIPSD* has biophysical functions, i.e., regulation of the actin cytoskeleton, receptor endocytosis, dendritic spine maintenance, and endosomal transport (Cho et al. 2013). The gene product of *IP6K2* converts inositol hexakisphosphate (InsP6) to diphosphoinositol pentakisphosphate (InsP7/PP-InsP5), mediates apoptosis through heat-shock protein 90, and influences mitophagy by attenuating the influence of PINK-1 in brain (Nagpal et al. 2022). Bioinformatic analyses of candidate genes from all 78 PD GWS loci (Nalls et al. 2019) identified the inositol phosphate biosynthetic pathway (Yu et al. 2024). The conjoint AN-PD SNP location immediately upstream of *IP6K2*, LD across the region, transcriptomic findings, and AN-neuropsychiatric associations point to complex regulation of multiple candidates at chr3p21.31 that requires additional analyses to understand potential shared mechanisms.

The convergence of neuroscience, genomics and insights into cross-disorder etiology suggest that cross-disorder research may enable acceleration of translational research for both groups of disorders. Based on AN’s estimates of polygenicity and discoverability, a larger number of causal SNPs influencing AN will be found when larger GWAS sample sizes become available. However, given existing GWAS sample sizes and differences in polygenicity and discoverability hampering gene discovery in AN relative to PD, discovery approaches using existing data will enrich AN gene discovery due to PD’s currently larger number of GWS loci, even while sample sizes in ED GWAS are increasing.

## Limitations

GBD 2019 described ascertainment of ED prevalence as challenging due to a historical focus on AN and BN; BED and OSFED prevalences were estimated using network meta-regression to obtain ratios of ED diagnoses, followed by Bayesian meta-regression to obtain disability weights followed by comorbidity correction, which produced estimates of ED DALYs with large uncertainty intervals. We acknowledge heterogeneity in disability in PD clinical variants not discussed here, and the uncertainty in population burden measures. Common variant heritability estimates are based on common SNPs and do not include rare or structural variants, two factors resulting in lower heritability estimates than twin-based estimates. Our review of genetic epidemiology looks back several decades and genomic epidemiology less than one decade; cross-disorder genomic relationships between EDs and PD will become more granular and specific as precision medicine defines disease subtypes. With the exceptions of global disability burden and meta-analyses of PD familiality, the literature reviewed, GWAS cited and data analyses performed used data from European ancestry individuals and/or high income countries with attendant limitations to generalizability.

## Conclusions

Our associated review and clinical study found shared neuropsychological and neurobiological features between AN and PD and increased familial risk of PD in ED families (Krueger, Angeline et al. submitted 2025). Our review of the epidemiologic and genetic epidemiologic literature identified similar population burdens between ED and PD, similar within-disorder familiality with greater twin, common variant heritability, and polygenicity estimates for AN than PD, but lower discoverability for AN. Inspection of AN GWS GWAS findings (SNP, gene and LD region) identified a single overlapping PD GWS GWAS finding, and, AN and PD GWAS cFDR association analysis identified a single conjointly associated genetic variant; the AN and PD SNPs found overlapping in the AN LD region and the cFDR SNP at chr3p21.31 were in LD with each other. As far as we are aware, this is the first in-depth description of genetic pleiotropy for AN and PD. Research and communication that leverages knowledge and engages investigators, reviewers and research sponsors from both communities may benefit scientific understanding and translational research for both disorders.

## Declarations

## Acknowledgements

Study participants, staff, researchers, IT groups, biorepositories and funders associated with: the Global Burden of Disease 2019 study; the Danish Psychiatric Central Research Register and the Danish Three Generation Study; the Departments of Psychiatry at the University of California Los Angeles and the University of Pittsburgh; published family studies in Parkinson’s Disease; the Swedish Twin Registry; the Eating Disorders Working Group of the Psychiatric Genomics Consortium (including the Anorexia Nervosa Genetics Initiative, Klarman Family Foundation, National Institute of Mental Health, Children’s Hospital of Philadelphia, Price Foundation Collaborative Group, Genetic Consortium for Anorexia Nervosa, Wellcome Trust Case-Control Consortium-3, and UK Biobank); the Brainstorm Consortium; the International Parkinson Disease Genomics Consortium (including PDGENE, the Web-Based Study of Parkinson’s Disease (PDWBS), 23andMe, 23andMe Research Team, Genentech, the Laboratory of Neurogenetics at the National Institute on Aging, Data Tecnica International, E-Scape Bio); and, the Cross-Disorder Group of the Psychiatric Genomics Consortium. The views expressed are those of the authors alone.

## Data availability statement

Parameter estimates and/or summary statistics from references cited, and from the NHGRI-EBI GWAS Catalog, the Psychiatric Genomics Consortium, Genentech, the TTAM Research Institute, and LDlink.

## Ethics approval statement

Ethical approval was granted by the UCSD Human Subjects Protection Program.

## Patient consent statement

All analyses performed were with summary statistics; no individual patient data was used.

## Permission to reproduce material from other sources

All sources of estimates and findings were cited.

---

**Supplementary Table 1.**
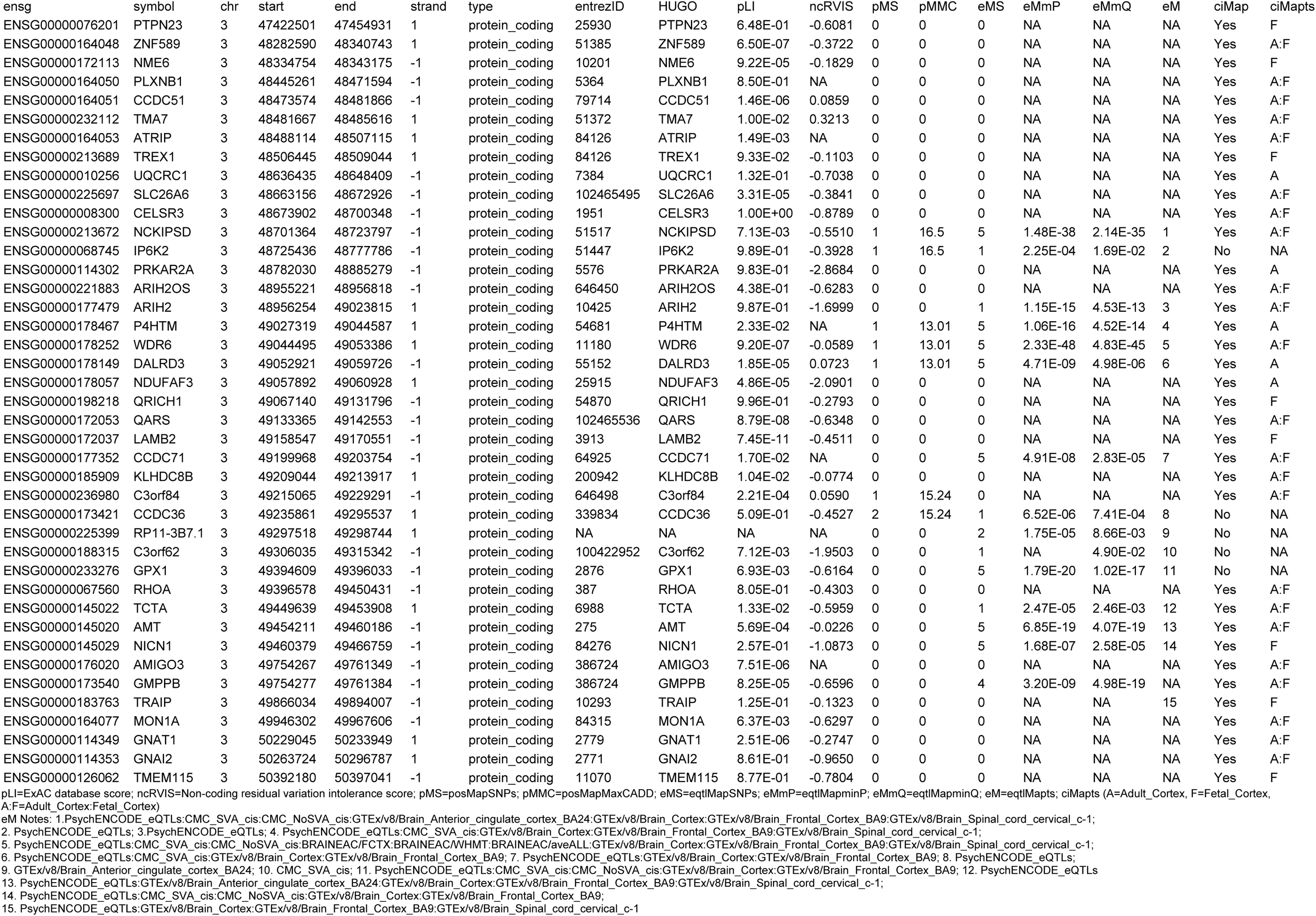
FUMA-mapped genes for rs1352420 via positional (pos), gene expression (eQTL) and chromatin interaction (ci) mapping.

## References

1. Anttila, V., B. Bulik-Sullivan, H. K. Finucane, et al. 2018. “Analysis of Shared Heritability in Common Disorders of the Brain.” Science (New York, N.Y.) 360 (6395). 10.1126/science.aap8757.

2. Bang, Lasse, Shahram Bahrami, Guy Hindley, et al. 2023. “Genome-Wide Analysis of Anorexia Nervosa and Major Psychiatric Disorders and Related Traits Reveals Genetic Overlap and Identifies Novel Risk Loci for Anorexia Nervosa.” Translational Psychiatry 13 (1): 291. 10.1038/s41398-023-02585-1.

3. Breton, Édith, and Tobias Kaufmann. 2025. “An Evolutionary Perspective on the Genetics of Anorexia Nervosa.” Translational Psychiatry 15 (1): 59. 10.1038/s41398-025-03270-1.

4. Bryois, Julien, Nathan G. Skene, Thomas Folkmann Hansen, et al. 2020. “Genetic Identification of Cell Types Underlying Brain Complex Traits Yields Insights into the Etiology of Parkinson’s Disease.” Nature Genetics 52 (5): 482–493. 10.1038/s41588-020-0610-9.

5. Bulik, Cynthia M., Patrick F. Sullivan, Federica Tozzi, Helena Furberg, Paul Lichtenstein, and Nancy L. Pedersen. 2006. “Prevalence, Heritability, and Prospective Risk Factors for Anorexia Nervosa.” Archives of General Psychiatry 63 (3): 305–312. 10.1001/archpsyc.63.3.305.

6. Cao, Ling-Xiao, Wee Lee Kong, Piu Chan, Wei Zhang, Margaret J. Morris, and Yue Huang. 2024. “Assessment Tools for Cognitive Performance in Parkinson’s Disease and Its Genetic Contributors.” Frontiers in Neurology 15 (June): 1413187. 10.3389/fneur.2024.1413187.

7. Cerezo, Maria, Elliot Sollis, Yue Ji, et al. 2025. “The NHGRI-EBI GWAS Catalog: Standards for Reusability, Sustainability and Diversity.” Nucleic Acids Research 53 (D1): D998–D1005. 10.1093/nar/gkae1070.

8. Chang, Diana, Mike A. Nalls, Ingileif B. Hallgrímsdóttir, et al. 2017. “A Meta-Analysis of Genome-Wide Association Studies Identifies 17 New Parkinson’s Disease Risk Loci.” Nature Genetics 49 (10): 1511–1516. 10.1038/ng.3955.

9. Chen, Dongze, Yi Zhou, Yali Zhang, Huatang Zeng, Liqun Wu, and Yuyang Liu. 2023. “Unraveling Shared Susceptibility Loci and Mendelian Genetic Associations Linking Educational Attainment with Multiple Neuropsychiatric Disorders.” Frontiers in Psychiatry 14: 1303430. 10.3389/fpsyt.2023.1303430.

10. Chevalier, Guenson, Lucas Udovin, Matilde Otero-Losada, et al. 2023. “Genetics of Neurogenic Orthostatic Hypotension in Parkinson’s Disease, Results from a Cross-Sectional in Silico Study.” Brain Sciences 13 (3). 10.3390/brainsci13030506.

11. Cho, In Ha, Dae Hwan Kim, Min-Jung Lee, Jeomil Bae, Kun Ho Lee, and Woo Keun Song. 2013. “SPIN90 Phosphorylation Modulates Spine Structure and Synaptic Function.” PloS One 8 (1): e54276. 10.1371/journal.pone.0054276.

12. Ding, Chenyu, Yuying Wu, Xiaoyong Chen, et al. 2022. “Global, Regional, and National Burden and Attributable Risk Factors of Neurological Disorders: The Global Burden of Disease Study 1990-2019.” Frontiers in Public Health 10 (November): 952161. 10.3389/fpubh.2022.952161.

13. Dorsey, E. Ray, Alexis Elbaz, Emma Nichols, et al. 2018. “Global, Regional, and National Burden of Parkinson’s Disease, 1990–2016: A Systematic Analysis for the Global Burden of Disease Study 2016.” Lancet Neurology 17 (11): 939–953. 10.1016/S1474-4422(18)30295-3.

14. Duncan, Laramie, Zeynep Yilmaz, Helena Gaspar, et al. 2017. “Significant Locus and Metabolic Genetic Correlations Revealed in Genome-Wide Association Study of Anorexia Nervosa.” The American Journal of Psychiatry 174 (9): 850–858. 10.1176/appi.ajp.2017.16121402.

15. Frank, Guido K. W., Megan E. Shott, and Marisa C. DeGuzman. 2019. “The Neurobiology of Eating Disorders.” Child and Adolescent Psychiatric Clinics of North America 28 (4): 629–640. 10.1016/j.chc.2019.05.007.

16. Goetz, Christopher G. 2011. “The History of Parkinson’s Disease: Early Clinical Descriptions and Neurological Therapies.” Cold Spring Harbor Perspectives in Medicine 1 (1): a008862. 10.1101/cshperspect.a008862.

17. Goldman, Samuel M., Kenneth Marek, Ruth Ottman, et al. 2019. “Concordance for Parkinson’s Disease in Twins: A 20-Year Update.” Annals of Neurology 85 (4): 600–605. 10.1002/ana.25441.

18. Hebebrand, Johannes, Jochen Seitz, and Abigail Matthews. 2025. “Considering Sufficient Weight Loss as a Prerequisite for Development of Anorexia Nervosa and Atypical Anorexia Nervosa.” The International Journal of Eating Disorders 58 (1): 162–167. 10.1002/eat.24313.

19. Holland, Dominic, Oleksandr Frei, Rahul Desikan, et al. 2020. “Beyond SNP Heritability: Polygenicity and Discoverability of Phenotypes Estimated with a Univariate Gaussian Mixture Model.” PLoS Genetics 16 (5): e1008612. 10.1371/journal.pgen.1008612.

20. Hübel, Christopher, Mohamed Abdulkadir, Moritz Herle, et al. 2021. “One Size Does Not Fit All. Genomics Differentiates among Anorexia Nervosa, Bulimia Nervosa, and Binge-Eating Disorder.” The International Journal of Eating Disorders 54 (5): 785–793. 10.1002/eat.23481.

21. International Parkinson Disease Genomics Consortium, Michael A. Nalls, Vincent Plagnol, et al. 2011. “Imputation of Sequence Variants for Identification of Genetic Risks for Parkinson’s Disease: A Meta-Analysis of Genome-Wide Association Studies.” Lancet 377 (9766): 641–649. 10.1016/S0140-6736(10)62345-8.

22. Kim, Jonggeol Jeffrey, Dan Vitale, Diego Véliz Otani, et al. 2024. “Multi-Ancestry Genome-Wide Association Meta-Analysis of Parkinson’s Disease.” Nature Genetics 56 (1): 27–36. 10.1038/s41588-023-01584-8.

23. Krueger, Angeline, Bergen, Andrew W, Litvan, Irene, et al. submitted 2025. “Eating Disorders and Parkinson’s Disease - 1: Comorbidities, Neurobiology and Family History.” Unpublished manuscript.

24. Levinson, Cheri A., Stephanie C. Zerwas, Leigh C. Brosof, et al. 2019. “Associations between Dimensions of Anorexia Nervosa and Obsessive-Compulsive Disorder: An Examination of Personality and Psychological Factors in Patients with Anorexia Nervosa.” European Eating Disorders Review: The Journal of the Eating Disorders Association 27 (2): 161–172. 10.1002/erv.2635.

25. Liu, Keke, Ruoyi Gao, Huining Kuang, Ranbo E, Chenyu Zhang, and Xin Guo. 2025. “Global, Regional, and National Burdens of Eating Disorder in Adolescents and Young Adults Aged 10-24 Years from 1990 to 2021: A Trend Analysis.” Journal of Affective Disorders 388 (119596): 119596. 10.1016/j.jad.2025.119596.

26. Lu, Zheng-An, Alexander Ploner, Andreas Birgegård, Eating Disorders Working Group of the Psychiatric Genomics Consortium, Cynthia M. Bulik, and Sarah E. Bergen. 2024. “Shared Genetic Architecture between Schizophrenia and Anorexia Nervosa: A Cross-Trait Genome-Wide Analysis.” Schizophrenia Bulletin 50 (5): 1255–1265. 10.1093/schbul/sbae087.

27. Machiela, Mitchell J., and Stephen J. Chanock. 2015. “LDlink: A Web-Based Application for Exploring Population-Specific Haplotype Structure and Linking Correlated Alleles of Possible Functional Variants.” Bioinformatics (Oxford, England) 31 (21): 3555–3557. 10.1093/bioinformatics/btv402.

28. Michel, Patrick P., Etienne C. Hirsch, and Stéphane Hunot. 2016. “Understanding Dopaminergic Cell Death Pathways in Parkinson Disease.” Neuron 90 (4): 675–691. 10.1016/j.neuron.2016.03.038.

29. Murray, Stuart B., Jaroslav Rokicki, Alina M. Sartorius, et al. 2023. “Brain-Based Gene Expression of Putative Risk Genes for Anorexia Nervosa.” Molecular Psychiatry 28 (6): 2612–2619. 10.1038/s41380-023-02110-2.

30. Nagpal, Latika, Michael D. Kornberg, and Solomon H. Snyder. 2022. “Inositol Hexakisphosphate Kinase-2 Non-Catalytically Regulates Mitophagy by Attenuating PINK1 Signaling.” Proceedings of the National Academy of Sciences of the United States of America 119 (14): e2121946119. 10.1073/pnas.2121946119.

31. Nalls, Mike A., Cornelis Blauwendraat, Costanza L. Vallerga, et al. 2019. “Identification of Novel Risk Loci, Causal Insights, and Heritable Risk for Parkinson’s Disease: A Meta-Analysis of Genome-Wide Association Studies.” Lancet Neurology 18 (12): 1091–1102. 10.1016/S1474-4422(19)30320-5.

32. Nalls, Mike A., Nathan Pankratz, Christina M. Lill, et al. 2014. “Large-Scale Meta-Analysis of Genome-Wide Association Data Identifies Six New Risk Loci for Parkinson’s Disease.” Nature Genetics 46 (9): 989–993. 10.1038/ng.3043.

33. Perez, Gerardo, Galt P. Barber, Anna Benet-Pages, et al. 2025. “The UCSC Genome Browser Database: 2025 Update.” Nucleic Acids Research 53 (D1): D1243–D1249. 10.1093/nar/gkae974.

34. Peyrot, Wouter J., and Alkes L. Price. 2021. “Identifying Loci with Different Allele Frequencies among Cases of Eight Psychiatric Disorders Using CC-GWAS.” Nature Genetics 53 (4): 445–454. 10.1038/s41588-021-00787-1.

35. Polymeropoulos, M. H., C. Lavedan, E. Leroy, et al. 1997. “Mutation in the Alpha-Synuclein Gene Identified in Families with Parkinson’s Disease.” Science (New York, N.Y.) 276 (5321): 2045–2047. 10.1126/science.276.5321.2045.

36. Santomauro, Damian F., Sarah Melen, Deborah Mitchison, Theo Vos, Harvey Whiteford, and Alize J. Ferrari. 2021. “The Hidden Burden of Eating Disorders: An Extension of Estimates from the Global Burden of Disease Study 2019.” The Lancet. Psychiatry 8 (4): 320–328. 10.1016/S2215-0366(21)00040-7.

37. Smeland, Olav B., Oleksandr Frei, Alexey Shadrin, et al. 2020. “Discovery of Shared Genomic Loci Using the Conditional False Discovery Rate Approach.” Human Genetics 139 (1): 85–94. 10.1007/s00439-019-02060-2.

38. Smeland, Olav B., Gleda Kutrolli, Shahram Bahrami, et al. 2023. “The Shared Genetic Risk Architecture of Neurological and Psychiatric Disorders: A Genome-Wide Analysis.” medRxiv : The Preprint Server for Health Sciences, ahead of print, September 26. 10.1101/2023.07.21.23292993.

39. Smeland, Olav B., Gleda Kutrolli, Shahram Bahrami, et al. 2025. “A Genome-Wide Analysis of the Shared Genetic Risk Architecture of Complex Neurological and Psychiatric Disorders.” Nature Neuroscience, ahead of print, November 11. 10.1038/s41593-025-02090-2.

40. Spillantini, M. G., M. L. Schmidt, V. M. Lee, J. Q. Trojanowski, R. Jakes, and M. Goedert. 1997. “Alpha-Synuclein in Lewy Bodies.” Nature 388 (6645): 839–840. 10.1038/42166.

41. Steinhausen, Hans-Christoph, Helle Jakobsen, Dorte Helenius, Povl Munk-Jørgensen, and Michael Strober. 2015. “A Nation-Wide Study of the Family Aggregation and Risk Factors in Anorexia Nervosa over Three Generations.” The International Journal of Eating Disorders 48 (1): 1–8. 10.1002/eat.22293.

42. Strober, M., R. Freeman, C. Lampert, J. Diamond, and W. Kaye. 2000. “Controlled Family Study of Anorexia Nervosa and Bulimia Nervosa: Evidence of Shared Liability and Transmission of Partial Syndromes.” The American Journal of Psychiatry 157 (3): 393–401. 10.1176/appi.ajp.157.3.393.

43. Thacker, Evan L., and Alberto Ascherio. 2008. “Familial Aggregation of Parkinson’s Disease: A Meta-Analysis.” Movement Disorders: Official Journal of the Movement Disorder Society 23 (8): 1174–1183. 10.1002/mds.22067.

44. Watanabe, Kyoko, Erdogan Taskesen, Arjen van Bochoven, and Danielle Posthuma. 2017. “Functional Mapping and Annotation of Genetic Associations with FUMA.” Nature Communications 8 (1): 1826. 10.1038/s41467-017-01261-5.

45. Watson, Hunna J., Zeynep Yilmaz, Laura M. Thornton, et al. 2019. “Genome-Wide Association Study Identifies Eight Risk Loci and Implicates Metabo-Psychiatric Origins for Anorexia Nervosa.” Nature Genetics 51 (8): 1207–1214. 10.1038/s41588-019-0439-2.

46. Wirdefeldt, Karin, Margaret Gatz, Chandra A. Reynolds, Carol A. Prescott, and Nancy L. Pedersen. 2011. “Heritability of Parkinson Disease in Swedish Twins: A Longitudinal Study.” Neurobiology of Aging 32 (10): 1923.e1–8. 10.1016/j.neurobiolaging.2011.02.017.

47. Ye, Hui, Laurie A. Robak, Meigen Yu, Matthew Cykowski, and Joshua M. Shulman. 2023. “Genetics and Pathogenesis of Parkinson’s Syndrome.” Annual Review of Pathology 18 (1): 95–121. 10.1146/annurev-pathmechdis-031521-034145.

48. Yilmaz, Zeynep, Matthew Halvorsen, Julien Bryois, et al. 2020. “Examination of the Shared Genetic Basis of Anorexia Nervosa and Obsessive-Compulsive Disorder.” Molecular Psychiatry 25 (9): 2036–2046. 10.1038/s41380-018-0115-4.

49. Yu, Eric, Roxanne Larivière, Rhalena A. Thomas, et al. 2024. “Machine Learning Nominates the Inositol Pathway and Novel Genes in Parkinson’s Disease.” Brain: A Journal of Neurology 147 (3): 887–899. 10.1093/brain/awad345.

